# Knowledge and practices related to menstruation among Lucknow college students in North India: results from cross-sectional survey

**DOI:** 10.1101/2021.02.10.21251460

**Authors:** Absar Ahmad, Surbhi G. Garg, Suman Gupta, Ruqayya Alvi

**Author notes:** Email id.

## Abstract

**Background:** Girls in many low and middle-income countries enter puberty with knowledge gaps and misconceptions about menstruation may lead to unsafe hygienic practices that increase health risk. Despite such importance, educated girls’ knowledge and hygienic practice towards menstruation are not well addressed in India. Consequently, the present study attempted to assess menstrual hygiene knowledge and practice among college students in Lucknow city in north India.

**Method:** An online college-based cross-sectional study design was employed in Lucknow, the capital of Uttar Pradesh in India. Data collection was carried out from September 11 to September 25, 2020, using a google form among undergraduate and Postgraduate students. All variables that were significant at bivariate level (at P-value < 0.05) were entered into multivariate analysis using a logistic regression model to control for confounding factors. In the final model, P-value of less than 0.05 was used as a base to identify factors having a statistically significant association with poor knowledge and hygiene practice at corresponding 95% confidence interval.

**Results:** More than half of students’ ages of menarche were between 12-15 years, and duration of menses flow was between 3-5 days. The most common premenstrual symptom was abdominal pain (67%) and back pain (50.5%). Majority of the students had first time discussed menstrual problems with their mothers (69.2%). Around 94% of students were currently using a sanitary pad as an absorbent. Regarding cleanliness, about (90.9%) girls clean their genitals after urinating during mensuration. Around 18.9% used medication during menstruation. Multivariate analyses reveal that students of Science and Technology (vs Commerce and Management Students) and monthly family income 50-100 thousand (vs < 25 thousands) were associated with good knowledge about menstruation. In contrast, students’ fathers were graduates (vs school educated); the occupation was ‘Other’(vs Farmer); living in a nuclear family(vs Joint family) and residing in urban areas(vs Rural) were less likely to have good menstruation knowledge. Besides, good hygiene practices are less likely to have with ‘Other’ Religion(vs Hindu), working mother(vs Housewife), monthly family income between 25 to 50 thousand(vs <25 thousands), and Nuclear family(vs Joint family) (p<0.05).

**Conclusions:** Most college students had poor knowledge but followed hygienic practices correctly. It demonstrates a need to design acceptable awareness creation and advocacy programs to improve college students’ knowledge during menstruation. Of all the sociodemographic factors, monthly income and types of family influenced students’ knowledge and practices related to menstruation.

## Introduction

Menstruation is a universal, normal, unique, and physiological phenomenon (1) that women and adolescent girls experience every month (2), but this topic has been taboo until a date in India (3). Girls in India hide their pad from gazing by the male at home. The shopkeeper always gives sanitary pads in the black polyethylene bag and wrap fully with newspaper. The issue is inadequately acknowledged and not received proper attention (4), so girls enter puberty with knowledge gaps and misconceptions in many low and middle-income countries. The adults around them, including parents and teachers, are ill-informed and uncomfortable discussing sexuality, reproduction, and menstruation (5). During menstruation, these sociocultural impositions make this phenomenon burdensome and an event that makes them feel fear, disgust, and shame (6). Among premenstrual, dysmenorrhea symptoms were the most frequent problems (7), linked to several misconceptions and practices (8) lead to poor menstrual hygiene (9). Its management is an important aspect of reproductive health, which, if not handled appropriately, can cause infections of the urinary tract, pelvic inflammatory diseases and vaginal thrush, as well as bad odor, soiled garments and, ultimately, shame, leading to infringement on the girls’ dignity (10)(11)(12).

It is advise that good menstrual hygiene practices are essential during menstruation; the practices include: 1) regular change of clothing and underwear; 2) change of hygienic pads every three to four hours; 3) daily showering, especially in instances of dysmenorrhea; 4) adequate washing of genitalia after each voiding of urine and/or feces; 5) continuing normal routine and daily activities (e.g. going to school, doing physical exercise), and 6) maintaining a balanced diet with plenty of fruits and vegetables rich in iron and calcium(13). Adolescent girls constitute a vulnerable group, not only concerning their social status but also to their health (1). They are often reluctant to discuss menstruation with their parents and usually hesitate to seek medical help regarding menstrual problems (14). This time for these students is crucial to prepare and adjust themselves to manage their menstrual bleeding safely and cleanly (15). College student’s life is very diverse than school student’s life. They make them independent, accountable, and robust. They also make decisions of their own that will transform their future. It is also an ideal time for girls to plan for their careers and prepare for a job competition. Managing the practical and psychological aspects of menstruation is difficult for these girls, affecting self-confidence, self-esteem and achieving the wider development goal of women’s empowerment (16). Menstrual also afflicts girl students absenteeism and academic performance (17). Risk factors associated with absenteeism include misconceptions about menstruation, insufficient and inadequate facilities at school or colleges, and family restriction (18).

Every year approximately 10 % of women worldwide are exposed to genital infections, including urinary tract infections and bacterial vaginosis, and 75 % of women have a history of a genital infection. Specifically, the common risk factor for vaginal infections include poor hygiene (19). A study in India reported exclusive use of disposable absorbents was low among young women aged 15-24 (37%) and varied substantially by caste, education, wealth, and residence (20). However, study of uses of sanitary pads particularly among college students in India were 80 % (21),but uses were in Pakistan(22) and Ghana (6) were 77.5 % and 100% respectively. A study in Uttar Pradesh state of India among 10-19-year girls reported about half of the girls did not have information or knowledge about menstruation. Less than one-quarter of them only followed proper hygiene practices (23). Another findings from a study in India suggest that rural school going girls who used old clothes or cloth boiled in water and dried before re-use suffered from genital infection (24). More than half of the women who did not use any hygienic method during menstruation suffered from vaginitis (25).

India is the second-largest country in the world, where we find state-wide variation in the use of menstrual absorbents, ranging from the highest in Mizoram (93.4%) to the lowest in Bihar (31%)(26). Further, these studies conducted among adolescents either using secondary data (20)(26) or school students (27)(8) in India and abroad (28)(6). These studies have shown inadequate knowledge of menstruation and poor menstrual hygiene practices among these population. Society considers persons with tertiary-educated proficient in diverse fields, their knowledge related to reproductive systems such as menstruation would be also high (6). Besides, there is little information about gynecological morbidity associated with menstrual problems in young populations (such as university students) (29). Considering the above scenario, this study was conducted among college students in Lucknow, India. This study assessed female undergraduate and post- graduate college student’s knowledge of various disciplines in Lucknow on menstruation and their menstrual hygiene practices. This study also evaluated determinants that could influence these students’ knowledge of menstruation. This study again tries to explore the taboo related to mensuration among college students. Policymakers and stakeholders will use the information obtained from this study to identify the awareness and practices of menstrual hygiene to provide information about menstruation and menstrual hygiene for college-going students in the study area.

## Data and Methods

Methods section of study were also published in previous study in 2021(30).

## Study design and methods

A descriptive cross-sectional online survey was performed in Lucknow using the google form platform.

## Study Setting

This study was carried out in Lucknow district of Uttar Pradesh, India.

## Participants

Undergraduate and Post Graduate students of any stream were eligible for the study.

## Sample size

Total 1439 participants took part in the survey. After removing 55 participants who quit the survey by clicking on the disagree button and 13 who did not satisfy inclusion criteria, the final sample comprised 1371 participants.

## Ethical consideration

Before the data collection, ethical clearance was obtained from the Institutional Ethical Committee of Career Institute of Medical Sciences and Hospital, Lucknow, with the reference number (Ref: PHARMA/SEP/2020/02). This work was done in collaboration with Shri Gurunanak Girls Degree College, Lucknow, and Career Institute of Medical Sciences and Hospital, Lucknow. The consent form was included in the online survey tool regarding their participation in the study. The study’s purpose was explained in the google form. The recipients had the full liberty to disagree and submit the survey form after knowing the purpose of the study. Personal information like mobile number, email id, and Name was not asked in the survey. Participants have also been informed as there will not any risk due to their participation, and the confidentiality of the collected information will be kept well. They were also informed that they have the right to refuse or withdraw their participation at any time they want, and no harm could be imposed towards them due to their refusal.

## Data collection

Data were collected by using an anonymous, internet based self- administered questionnaire that contained both open- and close-ended questions. The questionnaire was in English as well as Hindi, to get responses from Hindi medium students. The online questionnaires were conveniently distributed through emails, WhatsApp, Telegram, and other social media throughout Lucknow. The respondents’ social media were identified and recruited through a link and networking of all co-researchers and colleagues.

## Study variables

The predictor variable was chosen based on an extensive review of the literature on levels and factors associated with menstruation: Course of the study(Commerce and Management, Humanities, Medicine and Allied and Science and Technology),Grade(*Graduation and Post-graduation*), Age(<*19,20-22,23+*), Caste(*General, OBC, and SC/ST*), Religion(*Hindu, Muslim and Other*), Father’s education(*Illiterate or up to 12th, Graduate, and Postgraduate and higher*), Mother’s education(*Illiterate or up to 12th, Graduate and Postgraduate and higher*), Father’s occupation(*Farmer, Government employee, Self-employed/Businessman, Private employed, and Other*), Mother’s occupation(*Homemaker and working*), Monthly salary in Indian Rupees (< *25 thousand, 25 thousand to 50 thousand, 50 thousand to 1 lakh and 1 lakh above*), Type of family(*Joint and Nuclear*), and Place of residence(*Rural and Urban*).

### Outcome variables

Two outcome variables of the study are knowledge and practice related to menstruation. To measure girls’ knowledge about menstruation question was asked (i.e., what is the menstrual cycle; what the normal age of menarche is; organ of menstruation; and the normal duration of the menstrual cycle). Correct responses were coded as 1 otherwise 0 for analyses (see table 4). Similarly, to measure menstruation management’s good practice, five questions were asked and coded (See Table 5). Questions were the type of menstrual absorbent currently used, number of sanitary pad used per day during periods, cleaning genitals after urinating during mensuration, Types of pad wrap used for disposing of it, place at where the pad was disposed and bath during menstruation. For this study menstrual materials (sanitary pads or homemade cloth) changed four and more times a day is considered as good hygienic practices(31).

### Operational Definition

Total knowledge score varies from 0 to 4 and practice scores varies from 0 to 6. Median is considered as the cut-off point. A score below-median was considered as low or lacking knowledge and practices. Similarly, a score above median was considered as good or adequate knowledge and practices related to menstruation.

### Data Analysis

The data recorded in the spreadsheet were exported to SPSS Version 25. Initially, bivariate analysis was performed between dependent variable (Knowledge and practice of menstrual hygiene) and each of the independent variables (Socio- demographic variables). All variables that were significant at bivariate level (at P-value < 0.05) were entered into multivariate analysis using a logistic regression model to control for confounding factors. Their odds ratios (OR) at 95 % confidence intervals (CI) and P-values were obtained to identify important candidate variables for multivariate analysis.

## Results

From a total of 1439 students who participated in the online survey, 1371 completed the interview with a response rate of 95.2%. Table 1 shows the socio-economic and demographic profile of study participants. More than half (56.5%) were from the Commerce and Management course, followed by Humanities (33.6%). About 88% were undergraduate students, and the rest were postgraduate students. The mean (±SD) age of respondents was 20.06 (±1.78) years with a minimum and maximum age range of 16 and 30. Fifty-six percent of the sample students belonged to the general castes, 34% to other backward classes, and 10.3% to scheduled castes/ scheduled tribes. Most students surveyed were Hindu (86%), and 12% were Muslims. Forty-nine percent of students’ fathers were only up to school educated, one-third of fathers were graduate, and 17% postgraduate and above. Almost 60% of students’ mothers were up to school educated, 28% were graduate, and 12 % postgraduate and above. Most student’s fathers were self-employed (28%), followed by private organization employees (26%) and government employee (21%). However, 87% of student’s mothers were housewives, and 13 percent employed. Fifty-nine percent of students’ monthly family income was less than 25 thousand, two-third were living with Nuclear families, and three-fourths belonged to urban Lucknow.

**Table 1:**
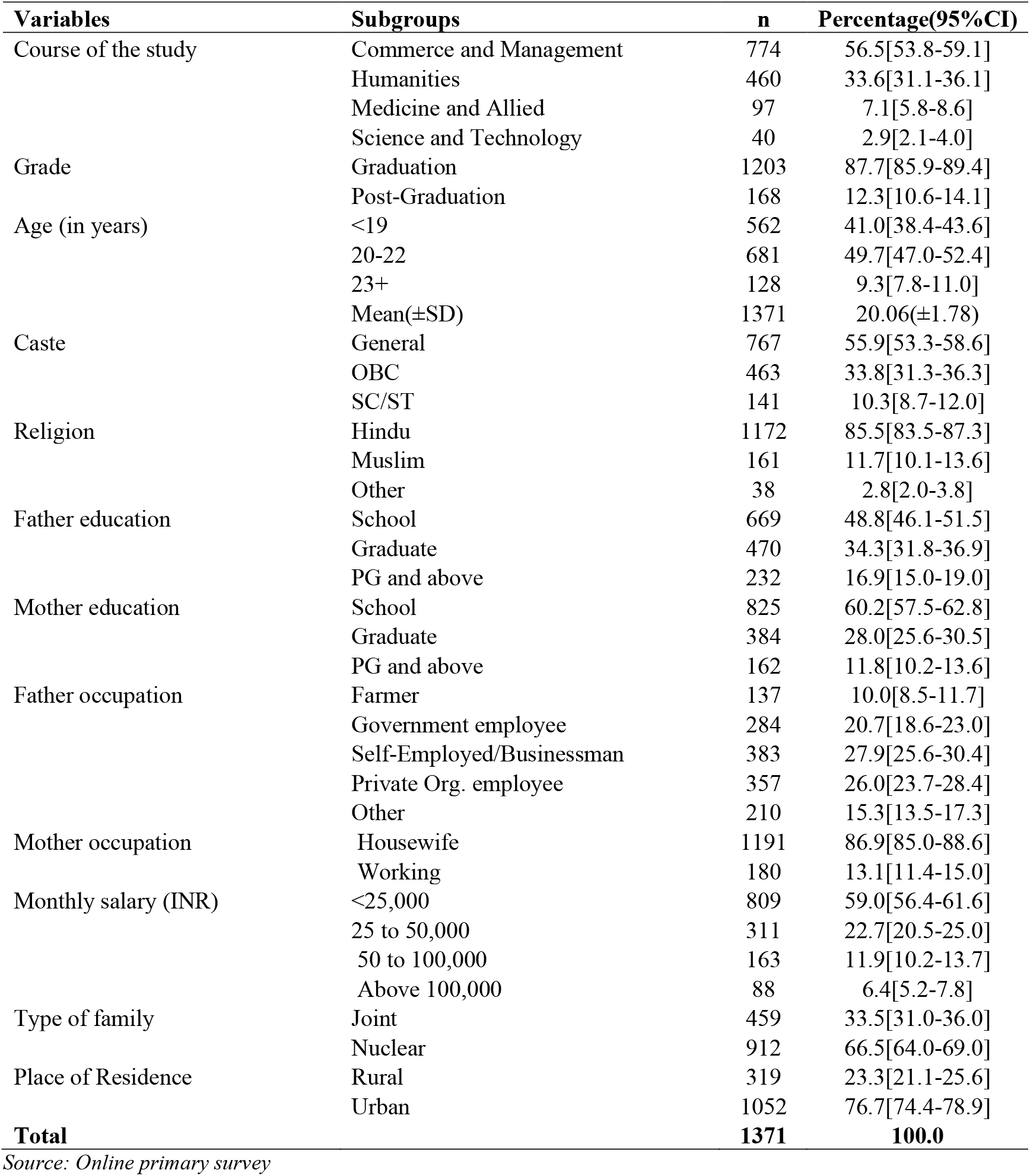
Socio-economic and demographic characteristics of college girls in Lucknow(n=1371)

| Variables | Subgroups | n | Percentage(95%CI) |
| --- | --- | --- | --- |
| Course of the study | Commerce and Management | 774 | 56.5[53.8-59.1] |
|  | Humanities | 460 | 33.6[31.1-36.1] |
|  | Medicine and Allied | 97 | 7.1[5.8-8.6] |
|  | Science and Technology | 40 | 2.9[2.1-4.0] |
| Grade | Graduation | 1203 | 87.7[85.9-89.4] |
|  | Post-Graduation | 168 | 12.3[10.6-14.1] |
| Age (in years) | <19 | 562 | 41.0[38.4-43.6] |
|  | 20-22 | 681 | 49.7[47.0-52.4] |
|  | 23+ | 128 | 9.3[7.8-11.0] |
| | Mean(±SD) | 1371 | 20.06(±1.78) |
| Caste | General | 767 | 55.9[53.3-58.6] |
|  | OBC | 463 | 33.8[31.3-36.3] |
|  | SC/ST | 141 | 10.3[8.7-12.0] |
| Religion | Hindu | 1172 | 85.5[83.5-87.3] |
|  | Muslim | 161 | 11.7[10.1-13.6] |
|  | Other | 38 | 2.8[2.0-3.8] |
| Father education | School | 669 | 48.8[46.1-51.5] |
|  | Graduate | 470 | 34.3[31.8-36.9] |
|  | PG and above | 232 | 16.9[15.0-19.0] |
| Mother education | School | 825 | 60.2[57.5-62.8] |
|  | Graduate | 384 | 28.0[25.6-30.5] |
|  | PG and above | 162 | 11.8[10.2-13.6] |
| Father occupation | Farmer | 137 | 10.0[8.5-11.7] |
|  | Government employee | 284 | 20.7[18.6-23.0] |
|  | Self-Employed/Businessman | 383 | 27.9[25.6-30.4] |
|  | Private Org. employee | 357 | 26.0[23.7-28.4] |
|  | Other | 210 | 15.3[13.5-17.3] |
| Mother occupation | Housewife | 1191 | 86.9[85.0-88.6] |
|  | Working | 180 | 13.1[11.4-15.0] |
| Monthly salary (INR) | <25,000 | 809 | 59.0[56.4-61.6] |
|  | 25 to 50,000 | 311 | 22.7[20.5-25.0] |
|  | 50 to 100,000 | 163 | 11.9[10.2-13.7] |
|  | Above 100,000 | 88 | 6.4[5.2-7.8] |
| Type of family | Joint | 459 | 33.5[31.0-36.0] |
|  | Nuclear | 912 | 66.5[64.0-69.0] |
| Place of Residence | Rural | 319 | 23.3[21.1-25.6] |
|  | Urban | 1052 | 76.7[74.4-78.9] |
| **Total** |  | **1371** | **100.0** |

Source: Online primary survey

Table 2 depicts the Obstetric and gynecological related characteristics of College girls in Lucknow. More than half, (57.8%) have experienced their menarche (onset of first menses) within the age range of 12–15 years, and 54.8 % had duration of menstrual flows of 3-5 days. Majority of college girls have experienced abdominal pain (67 %) during menstruation followed by backpain(50.5%), weakness(47.2%), irritability(41%), anorexia(24.3%), headache (17%), acne(17.1%), vomiting(10.3%), bloating(9.6%), insomnia(9.1%), constipation(7.9%) and breast pain(7.1%).

**Table 2:** Obstetric and gynecological related characteristics of College girls in Lucknow, Uttar Pradesh, India 2020 (n = 1371)

| Variables | Subgroup | n | Percentage(95%CI) |
| --- | --- | --- | --- |
| Age of Menarche | <12 Years | 197 | 14.4[12.6-16.3] |
|  | 12-15 Years | 792 | 57.8[55.1-60.4] |
|  | >15 Years | 266 | 19.4[17.3-21.6] |
|  | Don't Know | 116 | 8.5[7.0-10.1] |
| Duration of menses flow | <3 days | 249 | 18.2[16.2-20.3] |
|  | 3-5 days | 751 | 54.8[52.1-57.4] |
|  | >5 days | 371 | 27.1[24.7-29.5] |
| Symptom during menstruation | Acne | 234 | 17.1[15.1-19.2] |
|  | Vomiting | 141 | 10.3[8.70-12.0] |
|  | Diarrhoea | 73 | 5.3[4.20-6.60] |
|  | Insomnia | 125 | 9.1[7.60-10.80] |
|  | Breast pain | 97 | 7.1[5.80-8.60] |
|  | Bloating | 132 | 9.6[8.10-11.30] |
|  | Anorexia | 333 | 24.3[22.0-26.60] |
|  | Irritability | 563 | 41.1[38.40-43.70] |
|  | Backpain | 692 | 50.5[47.8-53.2] |
|  | Constipation | 108 | 7.9[6.50-9.40] |
|  | Headache | 233 | 17.0[1.50-19.1] |
|  | Weakness | 647 | 47.2[44.5-49.9] |
|  | Abdominal pain | 918 | 67.0[64.4-69.4] |
|  | At least one | 1371 | 100.0 |

Source: Online primary survey

Table 3 present the awareness among college girls of Menstruation at menarche and their reaction to first menstrual blood. Less than half (45.7%) of the students were aware of Menstruation before the onset of menarche. Mother is the first source of information for most of them, 69.2%). Friends (19.8%), teachers (18.4%), and sister (16.4%) were also the first source of information. Most of the respondents (30.6%) felt discomfort on seeing blood flowing from their genitals for the first time at menarche. However, mothers were the first persons the majority (70.8%) discussed menarche with rather than the least consulted father (0.2%). The majority (82.9%) expressed satisfaction with the level of education on Menstruation they got from the first persons they discussed their first menstrual episode.

**Table 3:**
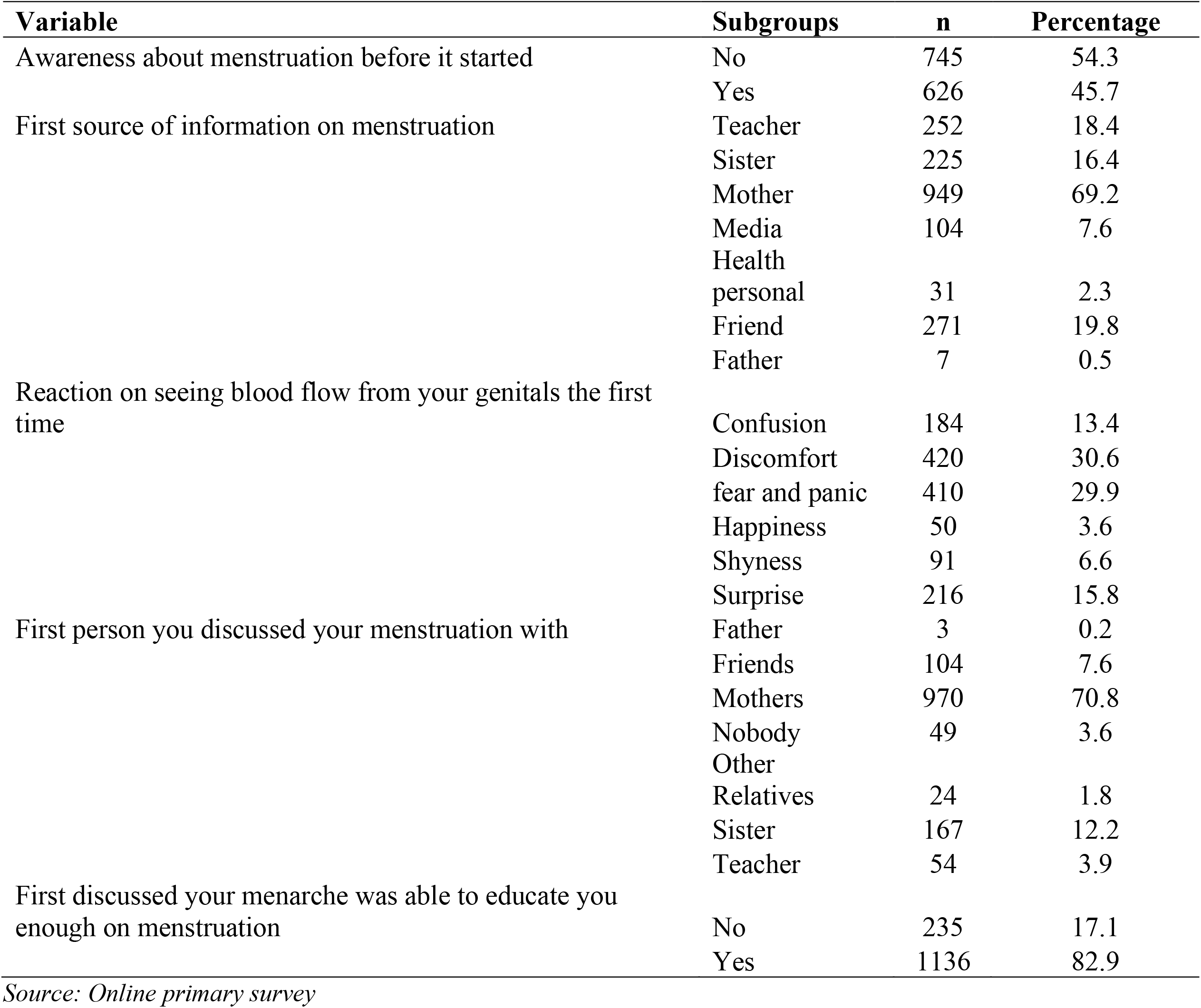
Awareness of menstruation of College girls in Lucknow, Uttar Pradesh, India 2020 (n = 1371)

Table 4 present the knowledge among college girls on menstruation in Lucknow. Solely 31 percent of college girls reported the definition of menstruation appropriately. Little lower than two-third (63.2%) of college girls correctly informed the typical age of menarche. While 65% of girls accurately reported organ of menstruation. Means Only 65 % of the girls knew that menstrual bleeding comes from the uterus. Only 64 % of girls correctly reported the usual duration of the menstruation cycle.

**Table 4:** Knowledge among college girls on menstruation in Lucknow, Uttar Pradesh, India 2020 (n = 1371)

| Variable | Knowledge | n | % | Scores |
| --- | --- | --- | --- | --- |
| What is the menstrual cycle |  |  |  |  |
| *Impure blood flows out* | Wrong | 871 | 63.5 | 0 |
| *Physiological and natural process* | Correct | 431 | 31.4 | 1 |
| *Some disease* | Wrong | 8 | 0.6 | 0 |
| *Curse of god* | Wrong | 9 | 0.7 | 0 |
| *Don't know* | Wrong | 52 | 3.8 | 0 |
| What is the normal age of menarche |  |  |  |  |
| *<10* | Wrong | 48 | 3.5 | 0 |
| *10-13* | Correct | 867 | 63.2 | 1 |
| *>13* | Wrong | 456 | 33.3 | 0 |
| Organ of menstruation |  |  |  |  |
| *Bladder* | Wrong | 315 | 23 | 0 |
| *Don't know* | Wrong | 109 | 8 | 0 |
| *Other* | Wrong | 19 | 1.4 | 0 |
| *Stomach* | Wrong | 37 | 2.7 | 0 |
| *Uterus* | Correct | 891 | 65 | 1 |
| What is the normal duration of the menstrual cycle |  |  |  |  |
| *25-30* | Correct | 885 | 64.6 | 1 |
| *Else* | Wrong | 486 | 35.4 | 0 |

Table 5 shows the menstrual hygiene practices of college students in Lucknow. Around 85% of students used the sanitary pad to absorb their menstrual blood during the first year after menarche, while 15% of students used reusable cloth. However, almost 94% of students are currently using the sanitary pad to manage their menstrual flow. The average sanitary pad used per day was thrice a day, and 91% clean their genitals after urinating during menstruation. Around 70% of students used paper to dispose of sanitary pads after wrapping it, while 29% used plastic bags. Approximately 9 % of students absent from at least one day during menstruation, and 19 % also used medication during menstruation. Around 15% skipped bathing on the first day of menses.

**Table 5:** Menstrual hygiene practices of College girls in Lucknow, Uttar Pradesh, India 2020 (n = 1371)

| Variable | Subgroups | N | Percentage | Scores |
| --- | --- | --- | --- | --- |
| Sanitary material used in first year of Menstruation | *Sanitary pads* | 1168 | 85.2 | NA |
|  | *Reusable cloth* | 207 | 15.1 |  |
|  | *Toilet tissue* | 33 | 2.4 |  |
|  | *Other* | 43 | 3.1 |  |
| Current sanitary material | *Sanitary Pad* | 1292 | 94.2 | 1 |
|  | *Other* | 79 | 5.8 | 0 |
| Number of Sanitary pads used per day | *Mean(±SD)* | 1371 | 2.91(±1.10) | 1 if ≥4 |
| Cleaning genitals after urinating during mensuration | *No* | 125 | 9.1 | 0 |
|  | *Yes* | 1246 | 90.9 | 1 |
| Types of pads wrap used for disposing of it | *Papers* | 957 | 69.8 | 2 |
|  | *Plastic bag* | 398 | 29.0 | 1 |
|  | *Not wrap* | 16 | 1.2 | 0 |
| Place at where pad was disposed | *Drain* | 13 | 0.9 | 0 |
|  | *Toilet* | 35 | 2.6 | 0 |
|  | *Open field* | 19 | 1.4 | 0 |
|  | *Dumped* | 91 | 6.6 | 1 |
|  | *Dustbin* | 1260 | 91.9 | 1 |
| Absent at least one day to college during menstruation | *Yes* | 123 | 9.0 | NA |
|  | *No* | 1248 | 91.0 |  |
| Used any medication for menstrual problems | *No* | 1112 | 81.1 | NA |
|  | *Yes* | 259 | 18.9 |  |
| Bath during menstruation | *Daily* | 1161 | 84.7 | 1 |
|  | *First day* | 16 | 1.2 | 0 |
|  | *Second day* | 149 | 10.9 | 0 |
|  | *Not take any time* | 45 | 3.3 | 0 |
| Total |  | 1371 | 100 |  |

Source: Online primary survey

Tables 6 exhibits the bivariate and multivariable relationship between the student’s knowledge and background characteristics. The multivariable analysis reveals the study’s course, father’s education, father’s occupation, monthly family income, type of family, and residence place. Students from Science and Technology course were five times more knowledgeable about menstruation than Commerce and Management students (AOR 5.17,95%CI: 2.274-11.752).

**Table 6:** Predictors of good knowledge related to menstruation of College girls in Lucknow, Uttar Pradesh, India 2020 (n = 1371)

| Variable | Subgroup | Poor | Good | n | Test, p value | COR (95%CI) | AOR (95% CI) |
| --- | --- | --- | --- | --- | --- | --- | --- |
| Course of the study | *Commerce and Management* | 462(59.7) | 312(40.3) | 774 | $\chi^2=57.58, <0.01^{**}$ | *Reference category* | *Reference category* |
|  | *Humanities* | 280(60.9) | 180(39.1) | 460 |  | 0.825(0.436-1.564) | 1.086(0.554-2.13) |
|  | *Medicine and Allied* | 20(20.6) | 77(79.4) | 97 |  | 0.786(0.410-1.506) | 1.049(0.529-2.078) |
|  | *Science and Technology* | 22(55.0) | 18(45.0) | 40 |  | 4.706(2.128-10.407) ** | 5.17(2.274-11.752) ** |
| Grade | *Graduation* | 686(57.0) | 517(43.0) | 1203 | $\chi^2=0.103, 0.74$ | NA | NA |
|  | *Post-Graduation* | 98(58.3) | 70(41.7) | 168 |  |  |  |
| Age | <19 | 345(61.4) | 217(38.6) | 562 | $\chi^2=6.96, 0.03^*$ | *Reference category* | *Reference category* |
|  | 20-22 | 368(54.0) | 313(46.0) | 681 |  | 0.783(0.532-1.155) | 1.14(0.749-1.735) |
|  | 23+ | 71(55.5) | 57(44.5) | 128 |  | 1.059(0.725-1.549) | 1.265(0.843-1.897) |
| Caste | *General* | 451(58.8) | 316(41.2) | 767 | $\chi^2=2.25, 0.32$ | NA | NA |
|  | *OBC* | 252(54.4) | 211(45.6) | 463 |  |  |  |
|  | *SC/ST* | 81(57.4) | 60(42.6) | 141 |  |  |  |
| Religion | *Hindu* | 671(57.3) | 501(42.7) | 1172 | $\chi^2=2.61, 0.27$ | NA | NA |
|  | *Muslim* | 87(54.0) | 74(46.0) | 161 |  |  |  |
|  | *Other* | 26(68.4) | 12(31.6) | 38 |  |  |  |
| Father education | *School* | 418(62.5) | 251(37.5) | 669 | $\chi^2=23.45, <0.01^{**}$ | *Reference category* | *Reference category* |
|  | *Graduate* | 263(56.0) | 207(44.0) | 470 |  | 0.479(0.354-0.649) ** | 0.634(0.426-0.944) * |
|  | *PG and above* | 103(44.4) | 129(55.6) | 232 |  | 0.628(0.458-0.863) ** | 0.695(0.482-1.002) |
| Mother education | *School* | 504(61.1) | 321(38.9) | 825 | $\chi^2=13.50, <0.01^{**}$ | *Reference category* | *Reference category* |
|  | *Graduate* | 201(52.3) | 183(47.7) | 384 |  | 0.606(0.432-0.850) ** | 1.141(0.729-1.786) |
|  | *PG and above* | 79(48.8) | 83(51.2) | 162 |  | 0.867(0.600-1.251) | 1.24(0.804-1.913) |
| Father occupation | *Farmer* | 77(56.2) | 60(43.8) | 137 | $\chi^2=12.85, 0.01^*$ | *Reference category* | *Reference category* |
|  | *Government employee* | 143(50.4) | 141(49.6) | 284 |  | 0.962(0.623-1.483) | 1.227(0.772-1.95) |
|  | *Self-Employed / Businessman* | 219(57.2) | 164(42.8) | 383 |  | 1.217(0.851-1.740) | 0.945(0.634-1.409) |
|  | *Private Org. employee* | 229(64.1) | 128(35.9) | 357 |  | 0.924(0.658-1.297) | 0.878(0.611-1.261) |
|  | *Other* | 116(55.2) | 94(44.8) | 210 |  | 0.690(0.487-0.976) * | 0.671(0.466-0.967) * |
| Mother occupation | *Housewife* | 702(58.9) | 489(41.1) | 1191 | $\chi^2=11.44, 0.01^*$ | *Reference category* | *Reference category* |
|  | *Working* | 82(45.6) | 98(54.4) | 180 |  | 0.583(0.425-0.799) ** | 0.777(0.54-1.117) |
| Monthly Income (INR) | <25,000 | 506(62.5) | 303(37.5) | 809 | $\chi^2=26.21, <0.01^{**}$ | Reference category | Reference category |
|  | 25 to 50,000 | 154(49.5) | 157(50.5) | 311 |  | 0.788(0.505-1.230) | 1.342(0.800-2.251) |
|  | 50 to 100,000 | 74(45.4) | 89(54.6) | 163 |  | 1.341(0.833-2.161) | 1.804(1.067-3.049) * |
|  | Above 100,000 | 50(56.8) | 38(43.2) | 88 |  | 1.583(0.938-2.668) | 1.688(0.957-2.978) |
| Type of family | Joint | 287(62.5) | 172(37.5) | 459 | $\chi^2=8.04, 0.01^*$ | Reference category | Reference category |
|  | Nuclear | 497(54.5) | 415(45.5) | 912 |  | 0.718(0.570-0.903) ** | 0.765(0.600-0.975) * |
| Place of Residence | Rural | 209(65.5) | 110(34.5) | 319 | $\chi^2=11.79, 0.01^*$ | Reference category | Reference category |
|  | Urban | 575(54.7) | 477(45.3) | 1052 |  | 0.634(0.489-0.824) ** | 0.701(0.527-0.933) * |
| Total |  | 784(57.20) | 587(42.80) | 1371 |  |  |  |

COR Crude Odds Ratio, AOR Adjusted Odds Ratio, \*P-value < 0.05, \*\*P < 0.01; Source: Online primary survey

Present study found contrast result from many studies. Students whose fathers graduated were significantly lower knowledge of menstruation than students whose fathers were school educated (AOR 0.634, 95%CI: 0.426-0.944). Likewise, students whose fathers’ occupations were ‘Other’ had lower knowledge of menstruation than students whose fathers were farmers (AOR 0.671, 95%CI:0.466-0.967). Students whose monthly family income was between 50 to 100 thousand were more knowledgeable about menstruation than those whose monthly income was less than 25 thousand (AOR 1.804, 95% CI:1.067-3.049). Students living in a Nuclear family were less knowledgeable than those living in a joint family (AOR 0.765 95% CI:0.600- 0.975). Furthermore, rural students were more knowledgeable than urban (AOR 0.701, 95% CI: 0.527- 0.933).This finding were also contrast of many Indian studies.

Table 7 presents the bivariate and multivariable relationship between sociodemographic characteristics and menstrual hygiene practices. The multivariable analysis reveals the factors associated with good hygiene practice of menstruation among students were religion, mother occupation, and family type. Other religion (Sikh and Christian) students were 88% less likely to had good menstrual hygiene practices than Hindu students [AOR = 0.11, 95 % CI: 0.048–0.273]. However, Students whose mothers were working women were 37% less likely to have good menstrual hygiene practices than students whose mothers were housewives [AOR= 0.63, 95 % CI: 0.42–0.953]. Nuclear family’s students were 34% less likely to have good menstrual hygiene practices than students from the joint family [AOR = 0.659, 95 % CI: 0.508–0.855].

**Table 7:** Predictors of good menstrual hygiene practices among College girls in Lucknow, Uttar Pradesh, India 2020 (n = 1371)

| Variable | Subgroup | Poor | Good | n | Test, p value | COR (95%CI) | AOR (95%CI) |
| --- | --- | --- | --- | --- | --- | --- | --- |
| Course of the study | Commerce and Management | 152(19.6) | 622(80.4) | 774 | $\chi^2=14.23, 0.003^*$ | Reference category | Reference category |
|  | Humanities | 126(27.4) | 334(72.6) | 460 |  | 1.18(0.554-2.548) | 0.996(0.426-2.332) |
|  | Medicine and Allied | 31(32.0) | 66(68.0) | 97 |  | 0.77(0.356-1.662) | 0.705(0.299-1.661) |
|  | Science and Technology | 9(22.5) | 31(77.5) | 40 |  | 0.618(0.263-1.45) | 0.661(0.258-1.698) |
| Grade | Graduation | 288(23.9) | 915(76.1) | 1203 | $\chi^2=3.06, 0.08$ | NA | NA |
|  | Post-Graduation | 30(17.9) | 138(82.1) | 168 |  |  |  |
| Age | <19 | 138(24.6) | 424(75.4) | 562 | $\chi^2=1.11, 0.57$ | NA | NA |
|  | 20-22 | 150(22.0) | 531(78.0) | 681 |  |  |  |
|  | 23+ | 30(23.4) | 98(76.6) | 128 |  |  |  |
| Caste | General | 182(23.7) | 585(76.3) | 767 | $\chi^2=3.39, 0.18$ | NA | NA |
|  | OBC | 112(24.2) | 351(75.8) | 463 |  |  |  |
|  | SC/ST | 24(17.0) | 117(83.0) | 141 |  |  |  |
| Religion | Hindu | 233(19.9) | 939(80.1) | 1172 | $\chi^2=72.81, <0.01^{**}$ | Reference category | Reference category |
|  | Muslim | 80(49.7) | 81(50.3) | 161 |  | 0.611(0.236-1.581) | 0.637(0.243-1.67) |
|  | Other | 5(13.2) | 33(86.8) | 38 |  | 0.153(0.057-0.413) | 0.144(0.052-0.399) ** |
| Father education | School | 173(25.9) | 496(74.1) | 669 | $\chi^2=5.76, 0.05$ | NA | NA |
|  | Graduate | 101(21.5) | 369(78.5) | 470 |  |  |  |
|  | PG and above | 44(19.0) | 188(81.0) | 232 |  |  |  |
| Mother education | School | 208(25.2) | 617(74.8) | 825 | $\chi^2=5.07, 0.07$ | NA | NA |
|  | *Graduate* | 80(20.8) | 304(79.2) | 384 |  |  |  |
|  | *PG and above* | 30(18.5) | 132(81.5) | 162 |  |  |  |
| Father occupation | *Farmer* | 35(25.5) | 102(74.5) | 137 | $\chi^2=4.15,0.38$ | NA | NA |
|  | *Government employee* | 60(21.1) | 224(78.9) | 284 |  |  |  |
|  | *Self-Employed/Businessman* | 100(26.1) | 283(73.9) | 383 |  |  |  |
|  | *Private Org. employee* | 74(20.7) | 283(79.3) | 357 |  |  |  |
|  | *Other* | 49(23.3) | 161(76.7) | 210 |  |  |  |
| Mother occupation | *Housewife* | 294(24.7) | 897(75.3) | 1191 | $\chi^2=11.31,0.001^*$ | *Reference category* | *Reference category* |
|  | *Working* | 24(13.3) | 156(86.7) | 180 |  | 0.47(0.299-0.736) ** | 0.49(0.304-0.791)** |
| Monthly Income (INR) | *<25,000* | 209(25.8) | 600(74.2) | 809 | $\chi^2=8.52,0.03^*$ | *Reference category* | *Reference category* |
|  | *25 to 50,000* | 61(19.6) | 250(80.4) | 311 |  | 0.543(0.300-0.982) * | 0.415(0.216-0.795)** |
|  | *50 to 100,000* | 34(20.9) | 129(79.1) | 163 |  | 0.775(0.410-1.465) | 0.534(0.269-1.058) |
|  | *Above 100,000* | 14(15.9) | 74(84.1) | 88 |  | 0.718(0.362-1.424) | 0.537(0.258-1.114) |
| Type of family | *Joint* | 127(27.7) | 332(72.3) | 459 | $\chi^2=7.75,0.005^*$ | *Reference category* | *Reference category* |
|  | *Nuclear* | 191(20.9) | 721(79.1) | 912 |  | 0.69(0.534-0.898) ** | 0.675(0.513-0.889)** |
| Place of Residence | *Rural* | 92(28.8) | 227(71.2) | 319 | $\chi^2=7.43,0.006^*$ | *Reference category* | *Reference category* |
|  | *Urban* | 226(21.5) | 826(78.5) | 1052 |  | 0.67(0.508-0.896) ** | 0.757(0.557-1.027) |
| **Total** |  | **318(23.2)** | **1053(76.8)** | **1371** |  |  |  |

COR Crude Odds Ratio, AOR Adjusted Odds Ratio, \*P-value < 0.05, \*\*P < 0.01; Source: Online primary survey

## Discussion

Menstruation and its management have become a globally recognized public health topic. Around the world, people and various stakeholders are mobilizing to bring attention and resources to address the menstrual-related shame, embarrassment, and taboos experienced by many girls in low and middle-income countries(32). Menstruation is the process through which every woman must come across throughout the reproductive years. Educated women manage menstrual hygiene in a better way, which positively impacts the personal welfare and health of a girl. Furthermore, these women are more likely to be healthier than uneducated women, which possibly ensures better health status and education for their children (33). Despite that, there is a limited study on university students’ menstrual knowledge and practices in India, and this study attempted to examine the menstruation knowledge and practices among them. The analysis introduced several significant insights.

More than 50% of students reported their age of menarche in the present study were in between 12-15 years. This was similar to the studies where the age of onset of menstruation or menarche were between 11-15 years (34)(35)(36)(37). The age of menarche varies by geographical region, race, ethnicity and other characteristics but ‘normally’ occurs in low income settings between the ages of 8 and 16 with a median of around aged 13(38)(39).

In present study every student experienced one or more symptom, respectively, in the premenstrual phase. Abdominal pain (menstrual cramps or dysmenorrhea) and back pain were most common symptoms related to menstruation in students, and these findings were concordant with other studies (40,41)(42)(36). Students reported that premenstrual symptoms made them excruciate and exhausted physically, mentally, and emotionally and counting days to end. Due to the high prevalence of these symptoms among students which increases long resting hours, negatively affected students daily living activities, disturbed social life and absenteeism from institutions (43)(44). Early diagnosis and knowledge about menstrual disturbances are essential because they help choose appropriate treatments, minimizing the disruptions or adverse effects on students’ lives (43).

In present study unfortunately, 54.3 % students were unaware of menstruation before menarche. The researchers observed that these percentages varied in various studies. The finding differs from 17% to 90 % (8)(34)(6) (45). Most of them were frightened, embarrassed, and disturbed due to ignorance about such a natural process (45).

The mother gave the first and primary source of information on menstruation to daughter. It was similar to the study done in Gujarat (46) and also revealed through meta-analysis (47) were the primary source of information was a mother. It has been also observed that the imperfect knowledge regarding menstruation among mothers and other adult family members is transferred to their young girls (48). Students reported knowledge like don’t bathe, don’t drink cold water, don’t eat curd, don’t play, and don’t exercise during mensuration passed this knowledge to children by mothers and other family members.

Most students’ reactions to seeing blood flow from their genitals the first time made them discomfort and created fear and panic. This study found results are congruent with other study in Ghana(6) and Pakistan(22) were the most students experienced fear and panic. Lack of information results in undue fear, anxiety, and wrong ideas in adolescents’ minds (49). It is also reported in India that women told nothing about menstruation until their first personal experience of it(50). It was also seen in some places that mothers do not teach their daughters about menstruation and hygiene maintenance during periods(8). Present study also found about 85% of students were used sanitary pads in the first year of menstruation, and currently, 94% were using sanitary pads which is higher than sanitary pad(37%) used by women aged 15-24 in India(20). The researchers observed the prevalence of currently using menstrual pad varied in various studies. The findings were ranges from 56% to 100 among college or university students (6)(22)(51)(21). In the present study, around 18.9% used medication during menstruation. However, in a study done in Malaysia among school girls, about 11.1 percent seek medical consultation for their menstrual disorders(35). Some women have had bad premenstrual symptoms on the days before their period that they can’t go about their usual activities. To cope with and manage common symptoms such as abdominal pain, women used medications.

In this study, multivariable analysis showed students from Science and Technology field were five times more knowledgeable about menstruation than Commerce and Management students. However, this association was not significant in the status of menstrual hygiene practice. Likewise, a study in Ghana found a significant association between the study’s course with menstrual knowledge (6). It might be because, in science, students have more exposure to this topic than commerce and management students.

Although knowledge of menstrual hygiene with religion was not significantly associated, the menstrual practice was significantly associated. The study found ‘Other’ religion students were less likely to follow good hygienic practices than Hindu religion students. In this study, the ‘Other’ religion consists of Sikh and Christian. Religion was a significant predictor of menstrual practice in studies in Ghana (6) and India(14). The present finding was the contrast of a study based on a large sample of women aged 15-24 where Christian women and those belonging to “other” religions were more likely than Hindu women to report exclusive uses of sanitary pads (20). In another study based on a large sample, the hygienic method was low among women aged 15-49 following Hindu and Muslim religion compared to women following Christianity, Sikhism, and other religion (52). As the sample size of the present study is very low among the reference group, this relation to be further explored with larger samples to develop any firm conclusion.

In this study, multivariable analysis showed that students whose fathers’ educational status was graduate were 36% less likely to had good knowledge about menstruation. However, the relationship was not significant for menstrual hygiene practices. Our finding is different from a study that stated good menstrual hygiene practice was high among schoolgirls whose fathers were educated (53). It may be because college students’ menstrual knowledge might be different from school student’s knowledge. This contradictory finding could be explained by the fact that the menstrual knowledge of students in college may independent of father education while in other studies the results were not significant (37)(54). So, the father’s education role might be changed from a previously established relation, which needs to be explored further using a different sample.

Furthermore, students whose fathers’ occupations were ‘Other’ had lower knowledge of menstruation than students whose fathers were farmers. However, the association between father occupation and menstruation were not significant in the present study. Other professions mostly include labour class or daily wagers. A study in Pakistan revealed that fathers’ occupation status is associated with students’ knowledge (54). However, a study in India shows girls living in households where the main occupation was labour had lower knowledge of menstruation than other works (23).

In multivariable analysis the knowledge level of menstrual hygiene doesn’t find significant association with mother occupation. However, the results were significant in case menstrual hygiene practices. Students whose mother is working less likely to follow good hygienic practices compare to those whose mother were housewife. Mothers’ employed status benefits children by improving family income, better disciplined work behaviour and better structure of family routines(55). Studies have shown that inadequate preparation for menarche creates a poor attitude toward menstruation and poor menstrual practices(56). Some students said that mothers do not teach kids because they think it makes children impudent, others expressed embarrassment of discussing it with their mothers, and some students believed that their families should not be involved in their sexual education(57). Those mothers who go to work and total time spent with the child has decreases (58). It was found that incidence of health issues was found to increase with increases in maternal employment as the number of hours spent with the child decrease(59).

In present study found that knowledge and practices related to menstruation were significantly associated with monthly income. In this study, we found that students whose monthly family income was 50 to 100 thousand were more knowledgeable about menstruation than those whose monthly income was less than 25 thousand. A study in India reported that girls living in households with poor economic status had lower knowledge of menstruation than better-off families(23). Students those family incomes were in between 25 to 50 thousand were less likely to follow good hygienic practices compare to students those family incomes were less than 25 thousand. due to low family income, men hesitate to give money for such costly products. So, in both cases, women have to compromise with their menstrual needs and personal hygiene(60).

In the present study found that knowledge and practices related to menstruation among college students were significantly associated with type of family. In the present study students living in a Nuclear family were less knowledgeable than those living in a joint family. However other study in India on knowledge level among students were not statistically significant with family type(61). During the discussions some of the girls stated that they were not aware about phenomenon of menstruation, but they observed that the females in their family keep themselves away from certain activities for two to three days every month(45). Students were belonging to nuclear family were less like to practice good hygienic practices compare to students belong to joint family. In Asian countries, and in many joint family systems, grandparents, and other nonworking family members fulfil the need for childcare–they take over the job of childcare when the mother is at work. This very important benefit (of readily available child support from the family members themselves) in joint families not only recognizes that the working mother is an important member of the family, but also provides her the necessary support to be able to perform her dual role efficiently(55).

Rural students were more knowledgeable than their counterparts (AOR 0.701, 95% CI: 0.527-0.933). The rural area of Lucknow has almost a rural-urban fringe, and students daily travel to the city for college. It shows twelve years of education have filled the knowledge gap related to menstruation. However, this relationship might be different when a study is done in another small city. Many students from rural backgrounds come to the city for under graduations and post-graduation and stay at a hostel or as paying guests in the city. Another situation was many colleges in Lucknow coming up in rural areas. These two situations might have altered the result of this study.

This study’s main strength is the inclusion of many students from different colleges in Lucknow. With such a large number and extensive participation, this study’s findings are difficult to be ignored. Despite that, there is a limitation in the study. The cross-sectional nature of the data could obscure the causal effect relationships of different factors, lacking qualitative data. Furthermore, we selected study subjects using a convenience sampling method to have shortcomings such as non-generalizability and selection bias. However, our study provided an insight into menstrual knowledge and practices in Indian college students and need further research for an in-depth understanding of the issue.

## Conclusion

Most college students had poor knowledge but followed hygienic practices correctly. It exhibits a need to design acceptable awareness creation and advocacy programs to improve college students’ learning regarding menstruation. Of all the sociodemographic factors, monthly income and family type influenced students’ knowledge and practices related to menstruation. Discussing the challenges faced by the girls during menstruation with family and friends and colleges can relieve anxiety and stress. Availability of adequate water, sanitary products, essential medicines, and privacy at home and the educational institutions can address the challenges. This paper reemphasizes the critical, urgent, and neglected need of providing correct knowledge to the community, including college students.

## Recommendation through students

It is recommended that students from 5th standard should make aware regarding menstruation. It should be cheap and accessible to all sections of girls. There should visit a yearly check-up for the students and medication should be prescribed for premenstrual symptoms. Male should also be informed about menstruation-related knowledge. The pad should be made more comfortable. Another option of menstrual hygiene management should also be introduced which should be environment friendly. Disposing bags should also be provided with sanitary pads. College should give off for at least one days when the pain was severe. A girl needs lots of love and care during her periods. It’s a natural process that every woman faces in this world to stop making them feel like a curse or insult. Every school, college, hospital, and institution should separate and individual small departments for girls where all related things about the menstrual cycle should be stored.

## Data Availability

All data has presented

## Acknowledgments

Our deepest gratitude also goes study participants, and faculties working at colleges in Lucknow for their dedicated cooperation.

## Authors contributions

AA, SG1, SG2 and RA conceived the topic, developed the proposal, and developing the study tool, manuscript preparation. AA took major roles in analysis. All authors gave final approval for this version of the manuscript to be considered for publication.

## Funding

The author(s) received no financial support for the research and/or authorship of this article.

## Availability of data and materials

All data has presented

## Competing interests

The authors declare that they have no competing interests

## Notes

### Competing Interest Statement

The authors have declared no competing interest.

### Author Declarations

ethical clearance was obtained from the Institutional Ethical Committee of Career Institute of Medical Sciences and Hospital, Lucknow, with the reference number (Ref: PHARMA/SEP/2020/02). This work was done in collaboration with Shri Gurunanak Girls Degree College, Lucknow, and Career Institute of Medical Sciences and Hospital, Lucknow.

